# A twelve-month projection to September 2022 of the Covid-19 epidemic in the UK using a Dynamic Causal Model

**DOI:** 10.1101/2021.10.04.21262827

**Authors:** Cam Bowie, Karl Friston

**Affiliations:** Retired Director of Public Health, Somerset, UK; Retired Professor of Community Health, College of Medicine, University of Malawi, Blantyre, Malawi; The Wellcome Centre for Human Neuroimaging, University College London, London, WC1N 3BG, UK

## Abstract

**Objectives:** Predicting the future UK Covid-19 epidemic allows other countries to compare their epidemic with one unfolding without public health measures except a vaccine programme.

**Methods:** A Dynamic Causal Model (DCM) is used to estimate the model parameters of the epidemic such as vaccine effectiveness and increased transmissibility of alpha and delta variants, the vaccine programme roll-out and changes in contact rates. The model predicts the future trends in infections, long-Covid, hospital admissions and deaths.

**Results:** Two dose vaccination given to 66% of the UK population prevents transmission following infection by 44%, serious illness by 86% and death by 93%. Despite this, with no other public health measures used, cases will increase from 37 million to 61 million, hospital admission from 536,000 to 684,000 and deaths from 136,000 to 142,000 over twelve months.

**Discussion:** Vaccination alone will not control the epidemic. Relaxation of mitigating public health measures carries several risks – overwhelming the health services, the creation of vaccine resistant variants and the economic cost of huge numbers of acute and chronic cases.

## Introduction

The recent abandonment of meaningful public health control measures in the UK in August 2021 provides a natural experiment offering a base-line control population for other countries to assess the effects of their own public health interventions. It does so by providing a fertile opportunity for Covid-19 to spread throughout the population for months to come. How many people will be infected in the next year? How many will suffer long-Covid? How many more people will die of Covid-19? How effective are the vaccines used in the UK - BNT162b2 (38%) and ChAdOx1 (61%)? What will be the likely re-infection rate as vaccine induced immunity wains?

Dynamic Causal modelling (DCM) is well-suited for predicting the effects of letting the virus sweep through the population because it combines conventional epidemiological models with behavioural modelling at the population level (i.e., it models and predicts both fluctuations in prevalence and contact rates) [1]. Its predictions can provide a base-line for other countries as they monitor the effects of their public health control efforts. “If you do nothing except vaccinate – this is what will happen”.

## Methods

### Dynamic causal models

An advantage of dynamic causal models (DCMs) is that the models are designed to continually assimilate data and modify model parameters, such as transmissibility of the virus, changes in social distancing and vaccine coverage—to accommodate changes in population dynamics and virus behaviour. The latest model (25^th^ September 2021) was used to explore the effect of increased ease of transmission of the Delta variant and the likely seasonal effect of the coming winter. Vaccine effectiveness with Delta and the curtailment of social distancing as well as the potential benefit of a successful FTTIS scheme were also incorporated into the model.

The model is fully described and a weekly dashboard provides up-to-date estimates and projections [2]. The software is freely available and can be used using datasets from other countries [3–7]. The following section describes the features of the model for non-modelling experts.

The model includes all the standard SEIR (susceptible, exposed, infected, removed) features of the commonly used models of infectious disease but in addition incorporates the interactions between the different variables. For example, people are more likely to stay at home if the prevalence is high or if they have not been immunised. These dependencies are estimated and only retained if they improve the ability of the model to account for the data. Having optimised the model and model parameters, one can then proceed with scenario modelling to evaluate the effect of interventions such as the influence of an enhanced FTTIS system on the epidemic.

Standard SEIR models depend on the choice of parameters, some of which are unknown empirically and must be guessed. Dynamic causal modelling is, by comparison, relatively assumption free. However, one must specify prior ranges for parameters (just like for SEIR models) but the DCM adjusts the parameters to fit the data in the most efficient and parsimonious way possible. Not only does the model provide estimates and projections of variables such as the death rate, the effective reproductive number, incidence, and prevalence but it also estimates of transmissibility, susceptibility, latent resistance, herd immunity, expected social distancing behaviour and vaccine effectiveness.

Two features provide insight into the way the model describes the interaction of the population and Covid-19. The first is the accuracy of the model in modelling the past stages of the epidemic. The second is the ability of the model to predict what will happen if we carry on as we have so far.

### Data sources and assumptions

The latest data from Public Health England (PHE) and the Covid-19 Infection Survey of the Office of National Statistics (ONS) [8,9] are used. It is assumed that mitigation efforts in schools will not take place, that lockdown will not be re-imposed and that no new more virulent variant will arrive despite our porous borders and minimal travel restrictions.

The changing transmissibility of the virus—as new variants emerge— is included in the model. Here, we assume that the alpha variant is 50% more transmissible than the original variants and the delta variant is 50% more transmissible than the alpha variant. These assumptions are based on published estimates [10– 13].

The model estimates the vaccine effectiveness in relation to transmission, pathogenicity, mortality and protection from infection. To do so it requires prior estimates which are derived from the ONS study and mortality from the PHE study (Table 1).

**Table 1.**
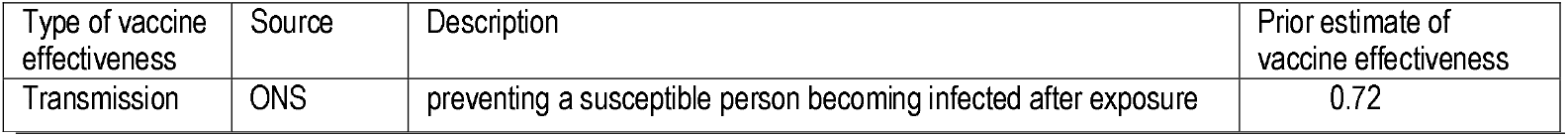
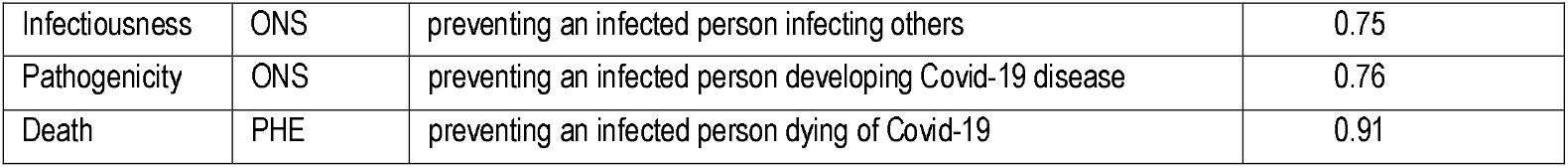
Prior estimates of vaccine effectiveness in UK up to August 2021.

| Type of vaccine effectiveness | Source | Description | Prior estimate of vaccine effectiveness |
| --- | --- | --- | --- |
| Transmission | ONS | preventing a susceptible person becoming infected after exposure | 0.72 |
| Infectiousness | ONS | preventing an infected person infecting others | 0.75 |
| Pathogenicity | ONS | preventing an infected person developing Covid-19 disease | 0.76 |
| Death | PHE | preventing an infected person dying of Covid-19 | 0.91 |

The mix of vaccines used in the UK up to 15^th^ September was ChAdOx1 - 53%, BNT162b2 – 45% and mRNA1273 (Moderna) – 3% [15]. The NHS vaccine scheme by 15^th^ September had provided two doses to 66% of the population [17].

Three scenarios are offered. The first (NPI1) provides the projections with baseline parameters. The second scenario (NPI2) improves the identify, test, trace, isolate and support (FTTIS) system from 30% to 50% successful. The third scenario (NPI3) increases the FTTIS system to 80%.

The latest ONS infection survey dated 4th July finds 835,000 people self-reporting post covid-19 syndrome (symptoms more than twelve weeks after presumed covid-19 infection) [19]. The infections would have occurred before 11^th^ April by which time the model estimated cumulative incidence of 24.9 million (3.36% of cases). This percentage is used to measure the trend in incidence of post covid-19 syndrome.

### Software

The figures in Figure 1 can be reproduced using annotated (MATLAB/Octave) code that is available as part of the free and open source academic software SPM. The routines are called by a demonstration script that can be invoked by DEM_COVID, DEM_COVID_X, DEM_COVID_T, DEM_COVID_I or DEM_COVID_LTLA at the MATLAB prompt. At the time of writing, these routines are available in the development version of the next SPM release. An archive of the relevant source code for each publication is available from figshare.

**Figure 1.**
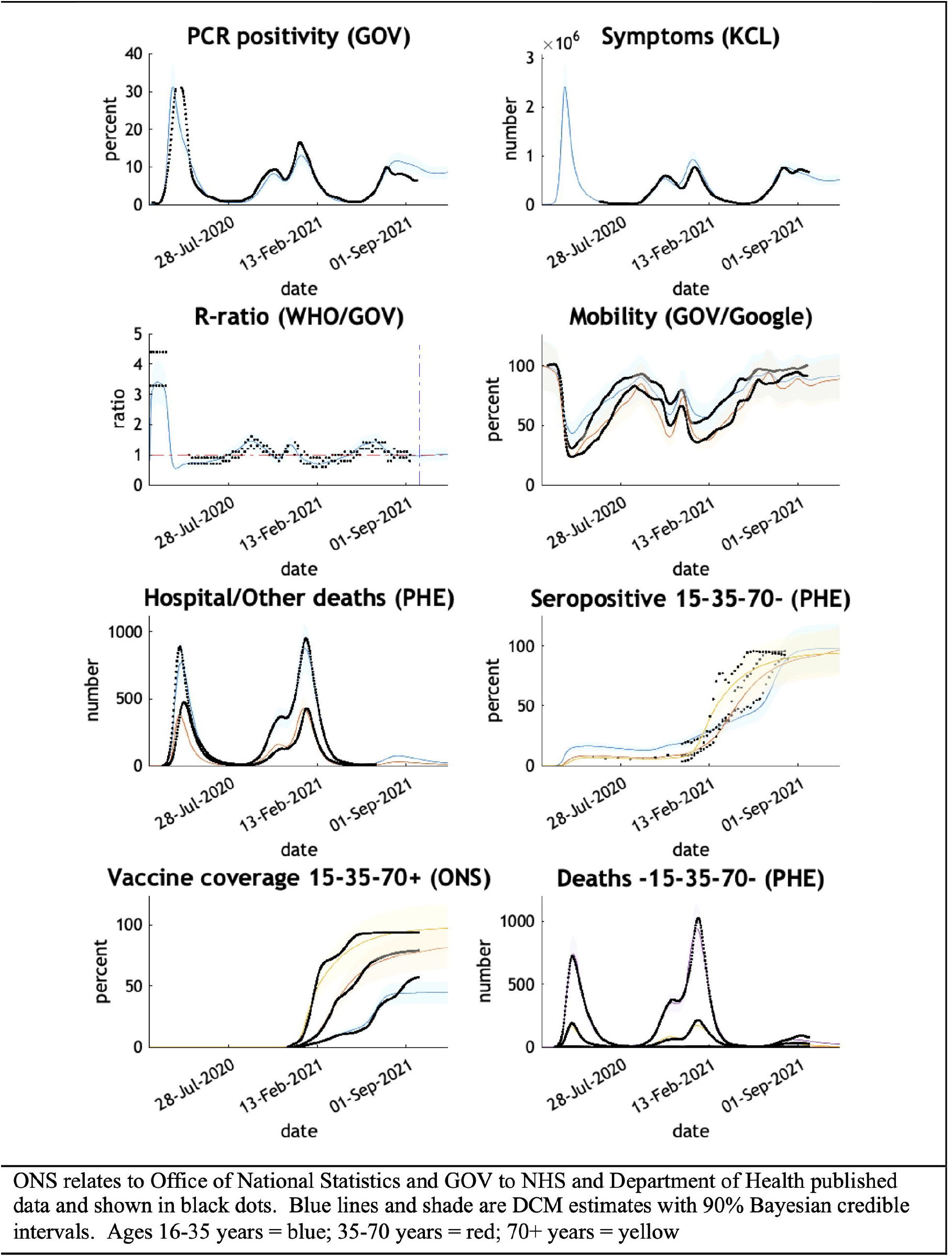
Comparing the actual with expected trends in eight measures of the Covid-19 epidemic UK – February 2020 to October 2021.

The remaining results in this paper can be reproduced using modified scripts found <u>here</u>.

The routine data used in the manuscripts are available from the COVID-19 Data Repository by the Center for Systems Science and Engineering (CSSE) at Johns Hopkins University, Coronavirus (COVID-19) UK Historical Data by Tom White and GOV.UK Coronavirus (COVID-19) in the UK. The CSV files must be available from the MATLAB path. The specific data on vaccine effectiveness are found in the ONS and PHE publications [14,20].

## Results

### The UK epidemic curve from February 2020 to September 2021

The chosen parameters adjusted by the model reproduce the epidemic curve and infection sequelae experienced by the UK up to now (Figure 1). The model estimates antibody immunity induced by Covid-19 infection and/or vaccine is lost in 284 days.

### Model predictions up to September 2022

The projections illustrate the depth and persistence of the future epidemic in the UK in terms of morbidity and mortality, transmission characteristics, testing capacity, hospital utilisation and disruption due to acute and chronic symptoms over the next twelve months if the government continues to withhold public health infection control measures (Figure 2).

**Figure 2.**
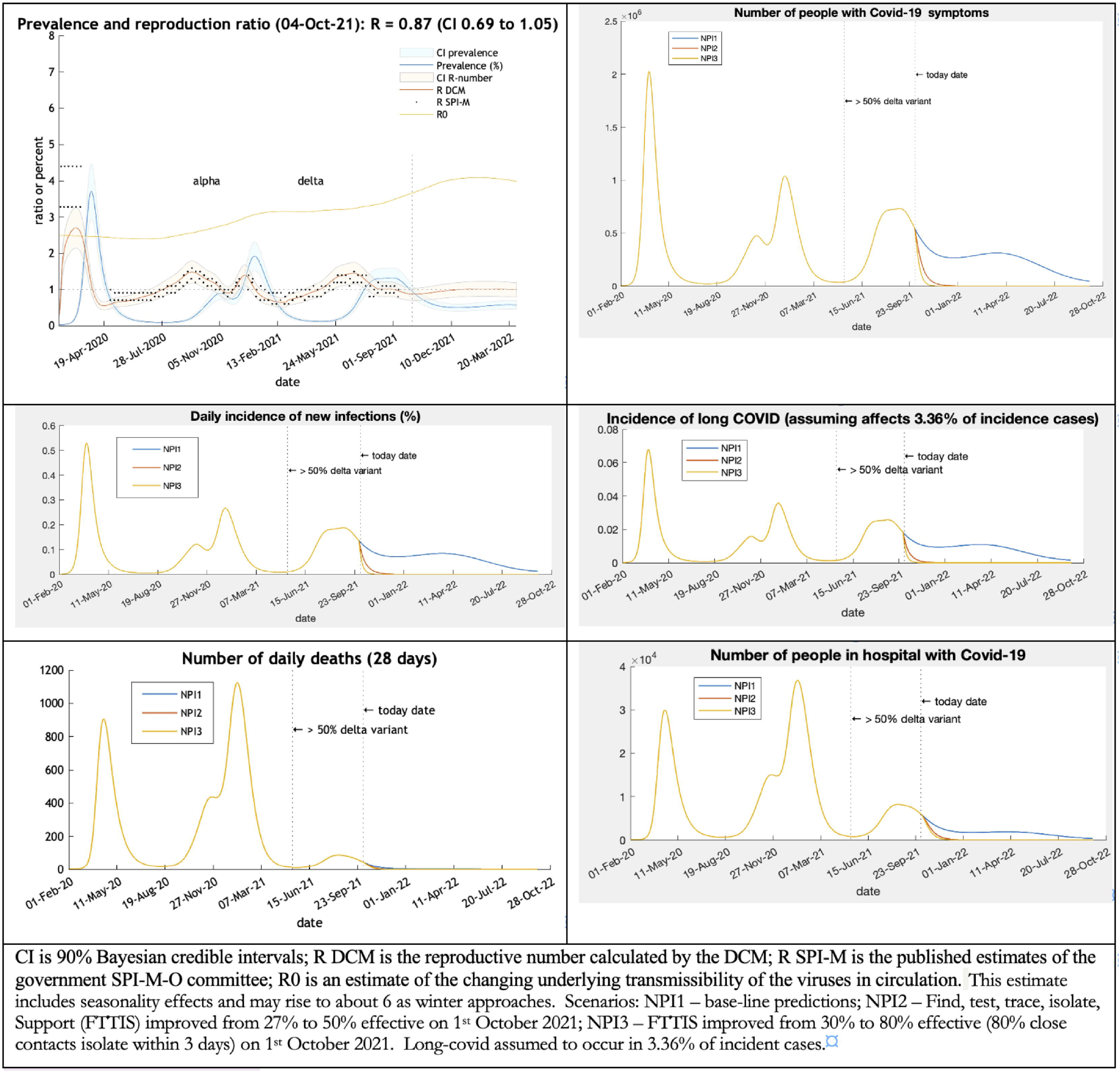
projections of the UK epidemic curve – incidence rate, daily confirmed cases, and incidence of long-Covid to March or September 2022.

### Vaccine effectiveness

The response in the UK to a prolonged wave of Covid-19 infections into the summer of 2022 is moderated by the high vaccine coverage. The vaccines reduce transmission by half compared to the original variants in circulation and pathogenicity for serious illness by 86% and deaths by 93% (Table 2). The effect for an individual is that two dose vaccination reduces the risk of infection from 100% to 37%, and death from 100% to 0.3% (Table 2).

**Table 2.** Vaccine effectiveness against Covid-19 Delta variant estimated by DCM model in UK in August 2021.

| Vaccine effectiveness with respect to Delta variant - September 2021 - UK | Parameter derived from published literature | Estimated by DCM model |
| --- | --- | --- |
| Preventing exposure to infection: | 75% | 63.2% (CI 61.5 to 64.9) |
| Preventing transmission following infection: | 72% | 44.2% (CI 40.9 to 47.4) |
| Preventing serious illness when symptomatic (age 15-34): | 76% | 85.5% (CI 85.0 to 85.9) |
| Preventing serious illness when symptomatic (age 35-70): |  | 85.6% (CI 85.2 to 86.1) |
| Preventing fatality when seriously ill: | 91% | 93.3% (CI 92.8 to 93.7) |
| **The reduction in risk from 100% after two doses of vaccine** |  |  |
| Relative risk of infection |  | 36.8% |
| Relative risk of mild illness |  | 32.8% |
| Relative risk of severe illness |  | 4.7% |
| Relative risk of fatality |  | 0.3% |
| CI = 90% Bayesian credible intervals. DCM = dynamic causal model |  |  |

### The long-term consequences

The trends in morbidity illustrate the consequences of allowing the epidemic to run in an uncontrolled manner through the community in the UK. The model can calculate its cumulative effect on cases numbers, deaths, tests and hospital admissions (Table 3). Tests double, cases increase by two-thirds, hospital admissions by a quarter and deaths by 5% over the coming 12 months. An effective FTTIS system in conjunction with the vaccine programme would more or less stop further cases, hospital admissions and deaths.

**Table 3.**
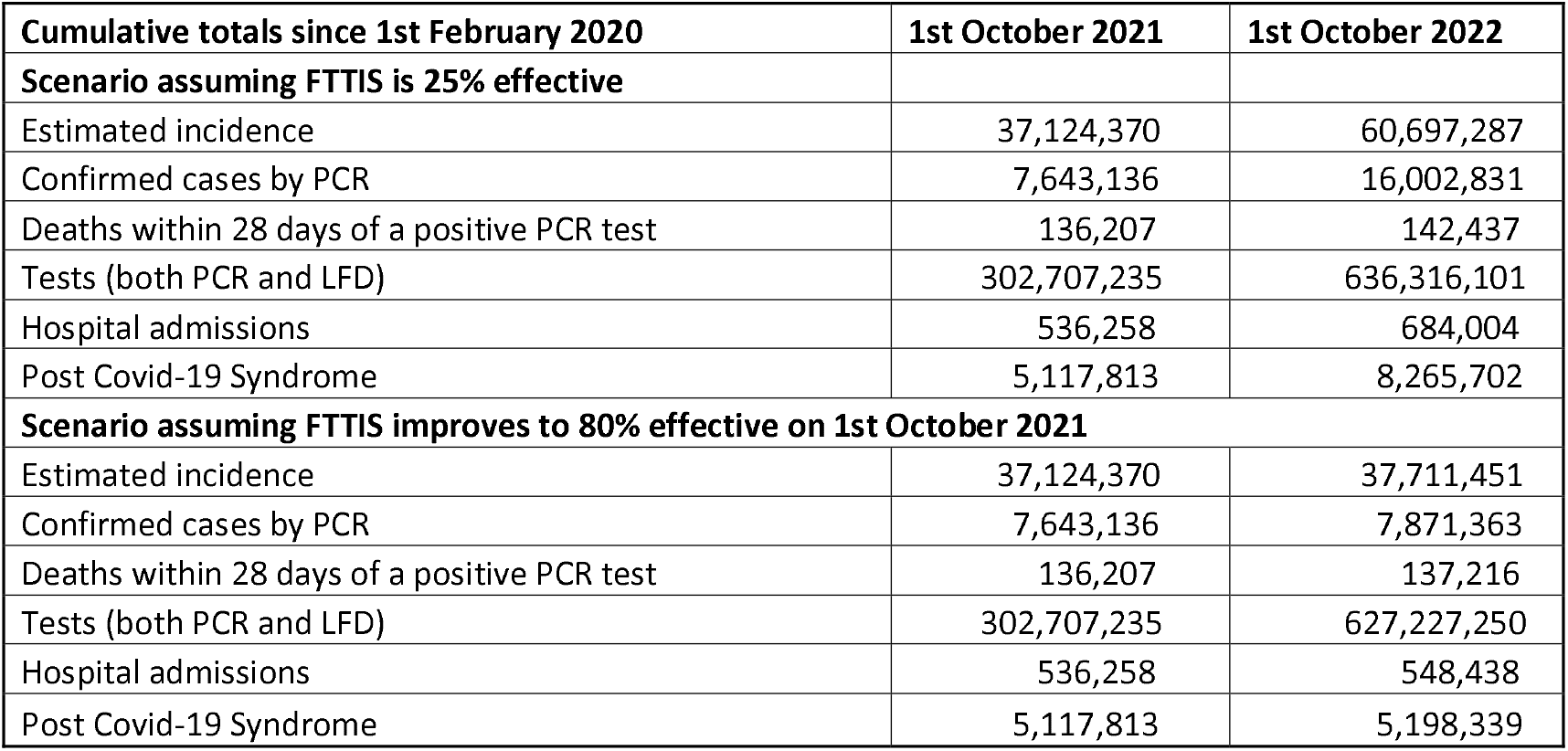
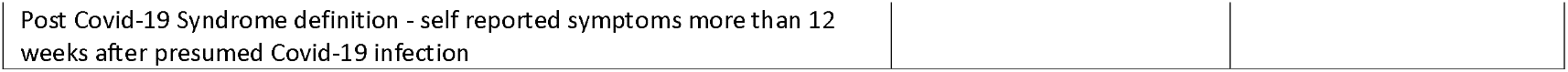
the cumulative effect of uncontrolled spread of Covid-19 in the UK – from 1^st^ February 2020 to 1^st^ October 2021 and to 1^st^ October 2022.

| Cumulative totals since 1st February 2020 | 1st October 2021 | 1st October 2022 |
| --- | --- | --- |
| **Scenario assuming FTTIS is 25% effective** |  |  |
| Estimated incidence | 37,124,370 | 60,697,287 |
| Confirmed cases by PCR | 7,643,136 | 16,002,831 |
| Deaths within 28 days of a positive PCR test | 136,207 | 142,437 |
| Tests (both PCR and LFD) | 302,707,235 | 636,316,101 |
| Hospital admissions | 536,258 | 684,004 |
| Post Covid-19 Syndrome | 5,117,813 | 8,265,702 |
| **Scenario assuming FTTIS improves to 80% effective on 1st October 2021** |  |  |
| Estimated incidence | 37,124,370 | 37,711,451 |
| Confirmed cases by PCR | 7,643,136 | 7,871,363 |
| Deaths within 28 days of a positive PCR test | 136,207 | 137,216 |
| Tests (both PCR and LFD) | 302,707,235 | 627,227,250 |
| Hospital admissions | 536,258 | 548,438 |
| Post Covid-19 Syndrome | 5,117,813 | 5,198,339 |

## Discussion

The UK provides a baseline of public health inactivity which can be used to compare public health controls chosen by other countries. The vaccines used in the UK seem to be extremely effective at reducing morbidity and mortality although not so effective at reducing transmission. Loss of immunity in 284 days as estimated by the model is similar to the results (extrapolated to 344 days) of the ONS study [14] (Table S4) suggests booster doses of vaccine may be required. The effect of a booster vaccine programme is not modelled.

Despite these very effective vaccines the UK can expect a huge further wave of Covid-19 infections resulting in over 3 million post Covid-19 Syndrome cases, 150,000 hospital admissions and 300 million additional tests. If other mitigating public health measures were employed to support the vaccine effects this further epidemic would be eliminated. Simply making contact tracing effective would achieve that possibility.

The size of the projected wave of infections provides fertile ground for new variants and the absence of border controls will allow new variants from other countries to invade the UK. A health service already exhausted will be rapidly overwhelmed. The lack of effective public health resources will be unable to respond to current or future variants. This means that the UK government may find it has to re-introduce restrictive lockdowns. The excellent scientific institutions in the UK will be able to monitor such features as genomic sequencing, vaccine efficacy, new vaccine trials. How long the population is prepared to be the guinea-pig remains to be seen.

Lessons for other countries are clear. Do not depend solely on vaccination. Relaxation of mitigating public health measures carries a number of risks – overwhelming the health service again, the creation of vaccine resistant variants and the economic cost of huge numbers of acute and chronic cases.

## Data Availability

DCM Covid-19 model available. Data used referenced and freely available on government websites.

https://www.fil.ion.ucl.ac.uk/spm/covid-19/

https://www.dropbox.com/sh/brf5oehf5zcyul6/AACBNbLe-Gcg-YeTqiGzisKha?dl=0

